# SISTR: Sinus and Inferior alveolar nerve Segmentation with Targeted Refinement on Cone Beam Computed Tomography images

**DOI:** 10.1101/2024.02.17.24301683

**Authors:** Laura Misrachi, Emma Covili, Hippolyte Mayard, Christian Alaka, Jérémy Rousseau, Willy Au

**Author notes:** These authors contributed equally to this work (Laura Misrachi, Emma Covili, Hippolyte Mayard).

## Abstract

**Background:** Accurate delineation of the maxillary sinus and inferior alveolar nerve (IAN) is crucial in dental implantology to prevent surgical complications. Manual segmentation from CBCT scans is labor-intensive and error-prone.

**Methods:** We introduce SISTR (Sinus and IAN Segmentation with Targeted Refinement), a deep learning framework for automated, high-resolution instance segmentation of oral cavity anatomies. SISTR operates in two stages: first, it predicts coarse segmentation and offset maps to anatomical regions, followed by clustering to identify region centroids. Subvolumes of individual anatomical instances are then extracted and processed by the model for fine structure segmentation. Our model was developed on the most diverse dataset to date for sinus and IAN segmentation, sourced from 11 dental clinics and 10 manufacturers (358 CBCTs for sinus, 499 for IAN).

**Results:** SISTR shows robust generalizability. It achieves strong segmentation performance on an external test set (98 sinus, 91 IAN CBCTs), reaching average DICE scores of 96.64% (95.38-97.60) for sinus and 83.43% (80.96-85.63) for IAN, representing a significant 10 percentage point improvement in Dice score for IAN compared to single-stage methods. Chamfer distances of 0.38 (0.24-0.60) mm for sinus and 0.88 (0.58-1.27) mm for IAN confirm its accuracy. Its inference time of 4 seconds per scan reduces time required for manual segmentation, which can take up to 28 minutes.

**Conclusions:** SISTR offers a fast, accurate, and efficient solution for the segmentation of critical anatomies in dental implantology, making it a valuable tool in digital dentistry.

**Plain text summary:** Accurately determining the locations of important structures such as the maxillary sinus and inferior alveolar nerve is crucial in dental implant surgery to avoid complications. The conventional method of manually mapping these areas from CBCT scans is time-consuming and prone to errors. To address this issue, we have developed SISTR, an AI-based framework that efficiently and accurately automates this process, trained on extensive datasets, sourced from 11 dental clinics and 10 manufacturers. It surpasses conventional methods by identifying anatomical regions within seconds. SISTR provides a rapid and accurate solution for high-resolution segmentation of critical anatomies in dental implantology, making it a valuable tool in digital dentistry.

## Introduction

The anatomy of the maxillofacial region is highly complex, making it a considerable challenge for medical imaging and computer-assisted diagnostic efforts. Key anatomical features within this complex region include the maxillary sinus and the Inferior Alveolar Nerve (IAN). Understanding the spatial relationships and characteristics of these anatomical entities is crucial for various clinical applications, such as the common surgical procedure of placing dental implants within the maxillary bone or the mandible. The precise delineation of boundaries, particularly for the IAN and sinuses, is crucial in implant placement applications to determine safe surgical margins, as highlighted by Jacobs et al.^1^. Accurate boundary identification is essential to ensure a minimum safety distance, such as the recommended 2mm between the IAN and implant apex, to prevent surgical complications^2^.

The maxillary sinus, a pyramid-shaped cavity within the maxillary bone^3^, is the largest air-filled chamber around the nasal cavity, constituting a substantial part of the maxilla. Located behind the cheeks, it communicates with the nasal cavity, influencing respiratory function. Variability in size and proximity to dental roots emphasizes the need for accurate segmentation in clinical decision-making^4^. When restoring the posterior maxilla with implants, practitioners often face the challenge of insufficient alveolar bone width and height. Elevating the sinus floor through either the lateral or transcrestal approach emerges as the most reliable and widely employed technique to address remaining bone deficiencies^56^.

In the mandible, significant elements include bilateral mandibular canals located below the premolars and molars, with openings called mandibular and mental foramen respectively^7^. Each canal contains the artery, vein, and the inferior alveolar nerve, a branch of the trigeminal nerve responsible for motor and sensory innervation to muscles, teeth, chin, and lower lip^89^. Accurate delineation of the inferior alveolar nerve is central in oral surgery to minimize the risk of nerve damage during procedures such as tooth extraction and dental implant placement.

3D Computed Tomography (CT) imaging techniques are commonly used to identify and diagnose these anatomical structures. Among them, Cone Beam Computed Tomography (CBCT) is preferred to Multi-Detector Computer Tomography (MDCT) as it has lower radiation exposure and cost^10^. CBCT enables high-resolution, three-dimensional imaging of anatomical structures, particularly in the craniofacial and dental regions, such as the maxillary sinus and the IAN. It is extensively used in dentomaxillofacial radiology^11^ for 3D diagnostics and surgical planning to address the challenge of precisely locating maxillary sinus and mandibular canals to prevent complications during surgical procedures.

The task of anatomical segmentation for the maxillary sinus and the IAN presents a dual challenge. Both the sinus and IAN manual segmentation by experts is laborious and time-consuming. It relies on the practitioner’s expertise, often exhibiting significant inter- and intra-observer variability^12,13,14^. In response to these challenges, the integration of Deep Learning techniques, specifically 3D Convolutional Neural Networks (CNNs), has emerged as a promising approach.

The segmentation of the maxillary sinus and the IAN has been a key focus in recent research initiatives. A variety of deep learning techniques have been developed for this purpose. For the maxillary sinus, methods vary from atlas-based approaches with fully convolutional networks, as demonstrated by Iwamoto et al.^15^, to Jung et al.’s first U-Net-like method^16^. More complex strategies include Morgan et al.’s two-stage model^17^, evaluated on 264 CBCTs from two manufacturers. Concurrently, the IAN witnessed a rapid integration of deep learning techniques, advancing beyond initial Statistical Shape Modelling (SSM) methods^18^, with Jaskari et al.^19^ introducing an innovative approach, though limited by coarse ground truth annotations. Kwak et al.^20^ expanded the field with SegNet^21^ and U-Net^22^ based models, but their training on a restricted dataset limits wider applicability. In 2022, Cipriano et al.^23^ released a substantial open-source CBCT dataset, mostly from a single manufacturer, involving a complex multi-stage training process. In parallel, Usman et al.^24^ enhanced IAN segmentation with a dual-stage architecture that localizes the Volume of Interest (VOI) first.

Nonetheless, these approaches primarily focus on datasets with limited diversity, sourced from a maximum of three different manufacturers, and are specific to segmenting single anatomical regions. Most use single-stage segmentation processes, which are often limited by high false positive rates and tend to overlook critical details in small objects like the IAN. Moreover, these methods face challenges in high-resolution segmentation due to memory and computational constraints. Cipriano et al.^25^ use complex training phases, including an intelligent sliding window method without precise object localization. Their multi-tier training process involves initial training on sparse data, followed by pretraining on synthetically generated dense annotations, and final finetuning on actual dense annotations, making integration into AI products challenging. The double stage approach of Usman et al.^24^, while effective for IAN and sinus due to their natural separability in the oral cavity, may not extend as well to other anatomical structures such as the teeth where distinct separability is lacking, limiting its broader applicability.

In the field of teeth segmentation, significant advances have been made, notably by Cui et al.^26^, who developed a pipeline for segmenting individual teeth and alveolar bones, assembling a large dataset of 4938 CBCTs from 15 centers. While their contribution is substantial, it does not address the specific regions this study focuses on. Additionally, their use of a Mask-RCNN 3D approach in Cui et al.^27^ for teeth segmentation, which involves a 3D Region Proposal Network (RPN) for handling multiple object localizations, contrasts with the more desirable approach of focusing on single anatomical object localization per instance. The latter is preferred for its superior computational efficiency, ease of implementation and maintenance —often requiring fewer hyperparameters—while being less prone to false positive detections^28^.

Reflecting on the challenges outlined earlier, we highlight three key improvement areas in dental AI: (1) the necessity of diverse data for generalization across dental practices, (2) the requirement for high-resolution segmentation in implant planning, and (3) the need for a unified segmentation method for oral anatomy (IAN, sinus, teeth).

Addressing these needs, we have developed a method influenced by the work of Cui et al.^26^ on individual teeth segmentation. Our method, called SISTR (Sinus and Inferior alveolar nerve Segmentation with Targeted Refinement) uses a two-stage training process featuring a 3D CNN model for segmenting the maxillary sinus and the IAN from CBCT images. This model combines segmentation and regression components. The first phase trains the network to regress offsets to centroids of each anatomical region, specifically the maxillary sinus and the IAN. The second phase starts with a clustering algorithm that identifies these centroids based on the offset maps generated earlier. These centroids then serve as focal points to crop focused sub-volumes that are fed to a network for precise structure segmentation. This is the Targeted Refinement phase. This method, appropriate for detailed dental anatomical segmentation, not only offers high precision in the second stage thanks to its localization module but also adapts well to broader applications in oral anatomy due to its simplicity. Our pipeline is trained on a diverse dataset from 11 dental clinics, including data from 10 different manufacturers, ensuring wide applicability in various dental settings.

This research presents our innovative two-stage pipeline, highlighting the advantages of a region proposal-based approach. To evaluate the generalization capabilities of our method, we conduct experiments using an external dataset comprising data from sources and centers not included in our development set. We also compare traditional architectures that segment the entire volume with our dual-stage method, focusing on localizing and then segmenting the area of interest. Our approach is substantially faster than semi-automatic annotation methods with manual interventions, marking a significant step forward in the precision and efficiency of dental AI applications.

## Methods

In the following section, we introduce SISTR: Sinus and Inferior alveolar nerve Segmentation with Targeted Refinement, our method designed for precise segmentation of CBCT images.

### SISTR architecture

Our proposed framework is designed to tackle several critical needs in the segmentation of anatomical structures within the oral cavity. It uses a unified method capable of distinguishing and identifying unique instances of each anatomy in the input CBCT, such as the left and right IAN, sinuses, and—anticipating future work—the classification of each tooth for precise implant planning. Essential to our approach is the ability for high-resolution segmentation, in line with the typical resolution range of 0.1 mm to 0.2 mm found in CBCTs scans used for implant planning. These scans often have large file sizes, ranging from 200 MB to 700 MB, with dimensions from (300, 300, 300) to (600, 600, 600), which presents significant challenges for GPU memory during processing and model training.

Moreover, the framework must be robust to various image types. While some CBCT scans offer a broad view of the entire buccal cavity, others may be truncated, focusing on specific anatomical zones. Precise segmentation is a key requirement, especially since minimizing false negatives is critical to avoid unintended intersection during implant placement. This precision is particularly challenging given the IAN’s narrow structure and its occasional invisibility in scan slices^14^.

Given the high native resolution of CBCT scans and the ensuing memory limitations, a single-stage segmentation method does not meet our needs. The literature supports a two-stage approach, starting with anatomical region localization, as more suitable given the technical and computational challenges.^27,19,24,30,17,26^. This method avoids the need to downsample the entire volume for GPU compatibility, which would compromise the segmentation detail necessary for precise implant planning.

In the first stage of our framework, we opt for a regression-based method to detect distinct instances through predictions of offset maps to the nearest anatomical region’s centroids. This strategy allows us to avoid the complexities and limitations of instance segmentation, which requires precise class-specific predictions, and semantic segmentation, which, while effective for distinguishing IAN and sinus regions due to their spatial separation, is inadequate for accurately segmenting closely positioned teeth without extensive post-processing for object instance separation^24^.

Inspired by the work of Cui et al.^26^ on teeth segmentation, our framework includes the segmentation of new regions, namely the IAN and Sinus, making it a comprehensive tool for anatomical region segmentation within the oral cavity.

Our model, as depicted in Figure 1, adopts a two-stage approach relying on target and refinement phases to segment anatomical structure. The first stage integrates a coarse segmentation branch alongside an anatomical region centroid offset regression branch, both layered on top of a Residual U-Net for voxel-level feature extraction. As this study aims to propose a method for segmenting anatomical regions in the oral cavity, a basic backbone was chosen for the model. This allows for a fair benchmark comparison against other models. The offset regression branch is tasked with predicting the distance from each voxel in the input volume to its nearest anatomical object’s centroid, in all three axes: X, Y and Z. A vectorized representation of such offsets maps is visible as the Vector Offsets label in Figure 1. Operating at a fixed resolution and volume size of (208, 208, 208), this initial stage is optimized for localization, where high resolution is not a prerequisite, thereby laying the groundwork for precise anatomical analysis. Segmentation and boundary predictions were addressed using a two-class classification paradigm at voxel-level.

**Figure 1.**
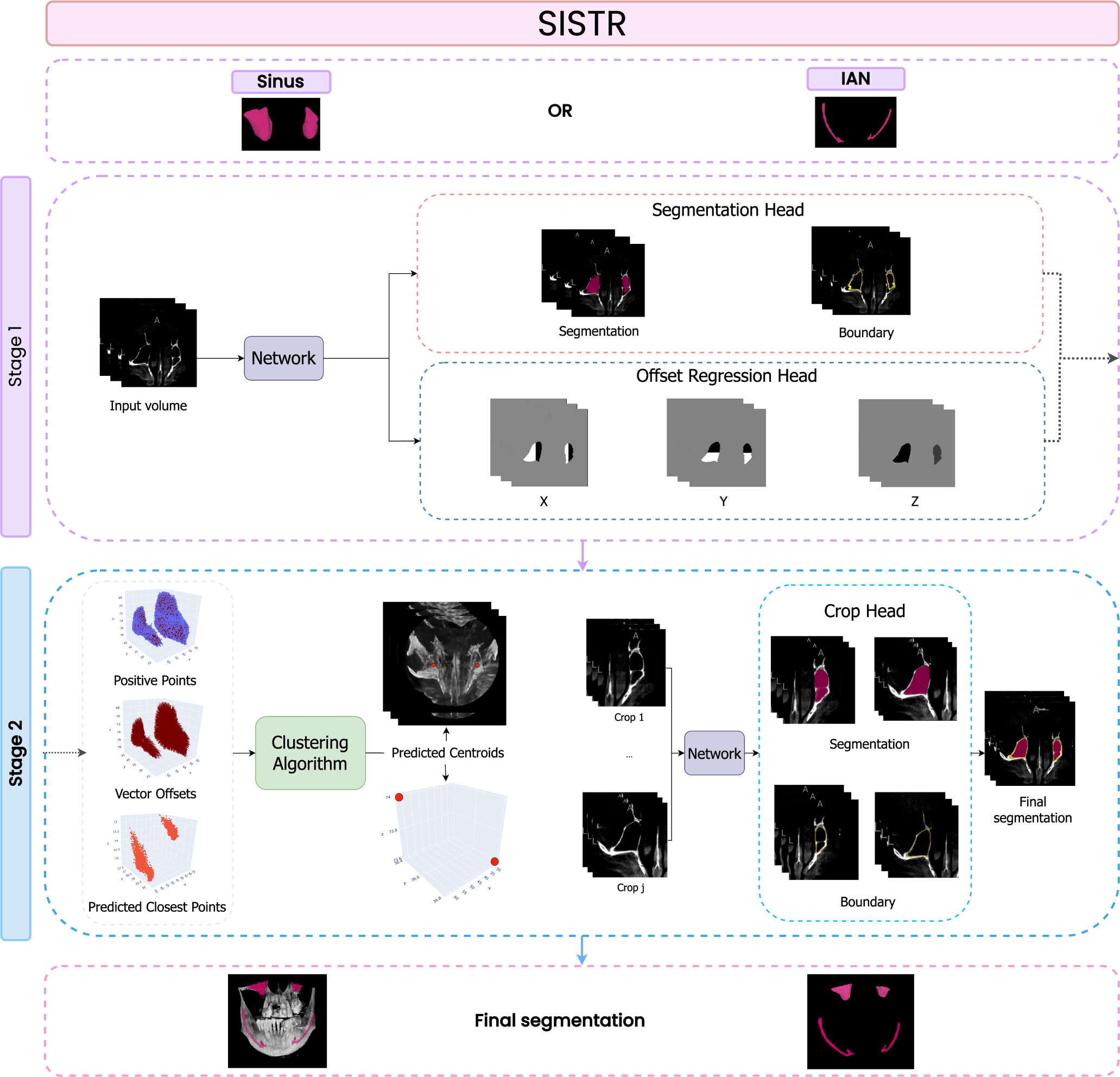
Architecture of the SISTR pipeline for Maxillary Sinus and IAN segmentation. This Figure illustrates the two-stage SISTR pipeline. Stage 1 involves a Segmentation Head for coarse anatomical segmentation and an Offset Regression Head for predicting voxel offsets to the nearest anatomical centroids along X, Y, and Z axes. Stage 2 employs these predictions, using a clustering algorithm^29^ to accurately locate centroids. Key visual elements in the Figure include Positive Points (indicating parts of the anatomy under consideration), Vector Offsets (showing the 3D point cloud of predicted offsets), and Predicted Closest Points (representing candidate centroids). The clustering algorithm identifies clusters among these points (details in Table 1, Supplementary Materials). The Crop Head then processes these centroids to extract anatomy-centered cropped volumes for detailed segmentation. The pipeline operates at low resolution in Stage 1 and can handle high resolution in Stage 2, particularly in the Crop Head for refined segmentation. The final segmentation is achieved by remapping cropped volumes segmentation to their native resolution. The depicted scheme is based on sinus anatomy but is adaptable for IAN segmentation.

**Table 1.**
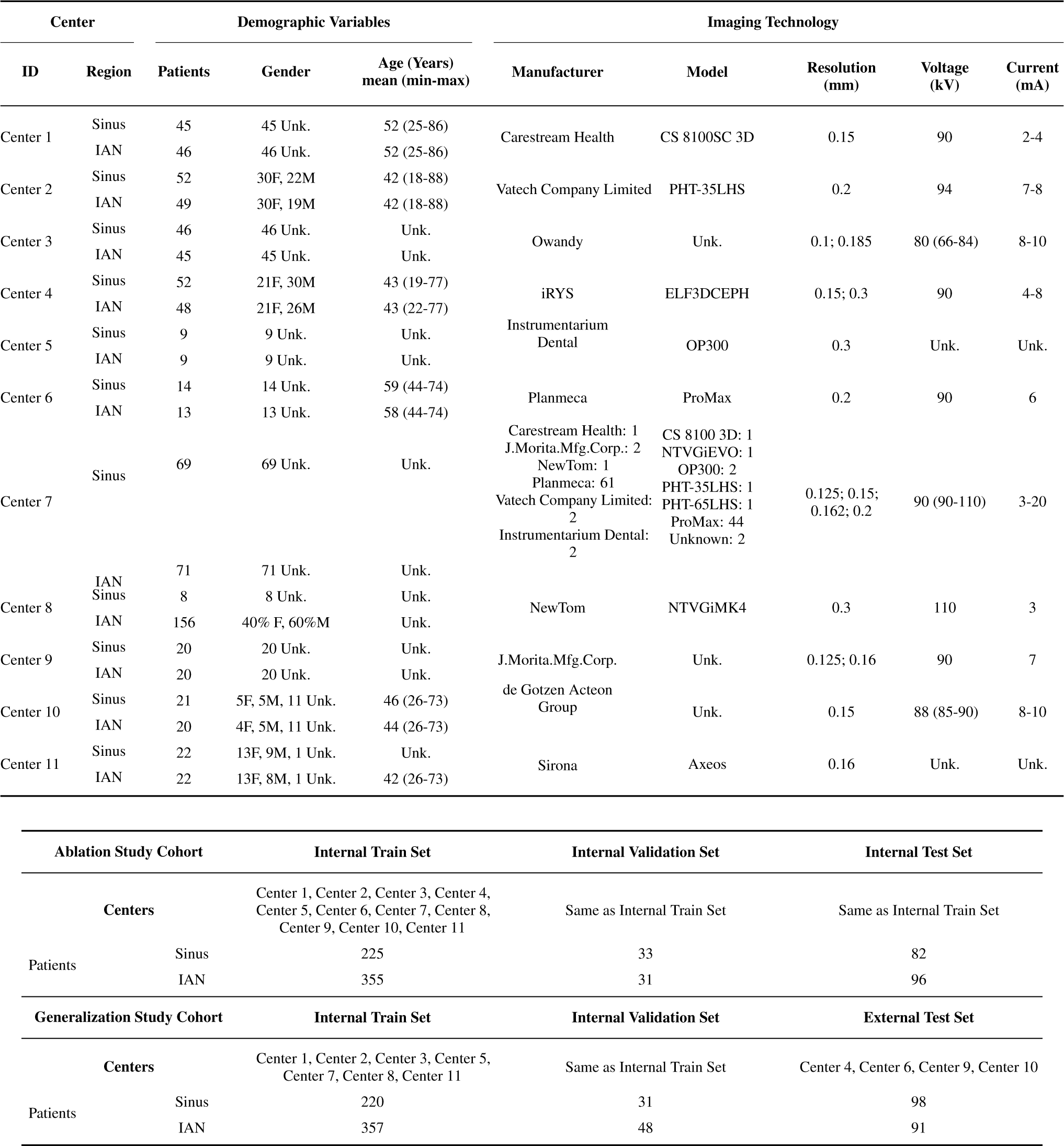
Dataset characteristics from 11 centers: imaging protocols, patient Demographics, and equipment Specifications. This table outlines the dataset composition from each center, including patient demographics (gender and age, where available), and details on CBCT equipment such as manufacturer, model, resolution, tube voltage, and current. Additionally, it presents the distribution of our study cohorts in both the development (Internal Train Set and Internal Validation Set) and testing phases (Internal Test Set and External Test Set), covering the Ablation Study and Generalization Study cohorts. ‘Unk.’ indicates unknown information within the dataset.

The second stage starts with a clustering algorithm^29^ to identify the centroids of each 3D anatomical structure. Following the method used by Cui et al.^26^, this approach focuses on clustering dense points while maintaining significant distances between these clusters. The clustering algorithm, whose hyperparameters are specified in Table 1 of the Supplementary Materials, uses the predicted offset maps (referred to as Vector Offsets in Figure 1) along the X, Y, and Z axes, along with the initial coarse segmentation, as its input sources. It produces a set of potential centroid candidates for the anatomical structure being examined. With these centroids, the model then generates a cropped volume centered around the target anatomical object. The dimensions of this cropped volume are predefined based on the known sizes of the anatomical structures, leveraging the accurate scaling of CBCT images. This enables SISTR to maintain the original resolution of the input CBCT volume when analyzing these smaller, defined sub-volumes, which can be fully processed by GPUs for high-resolution segmentation. Given the previously outlined need for accurate boundary delineation in implant placements, and considering that high segmentation accuracy alone is not always indicative of accurate boundary identification, our framework incorporates a boundary prediction feature in all its segmentation heads, including both the Segmentation Head and Crop Head.

The segmentation and the delineation of boundaries within this volume are carried out using a 3D Res-U-Net architecture. Detailed descriptions and illustrations of the model architectures are available in Figures 1 and 2 of the Supplementary Materials.

**Figure 2.**
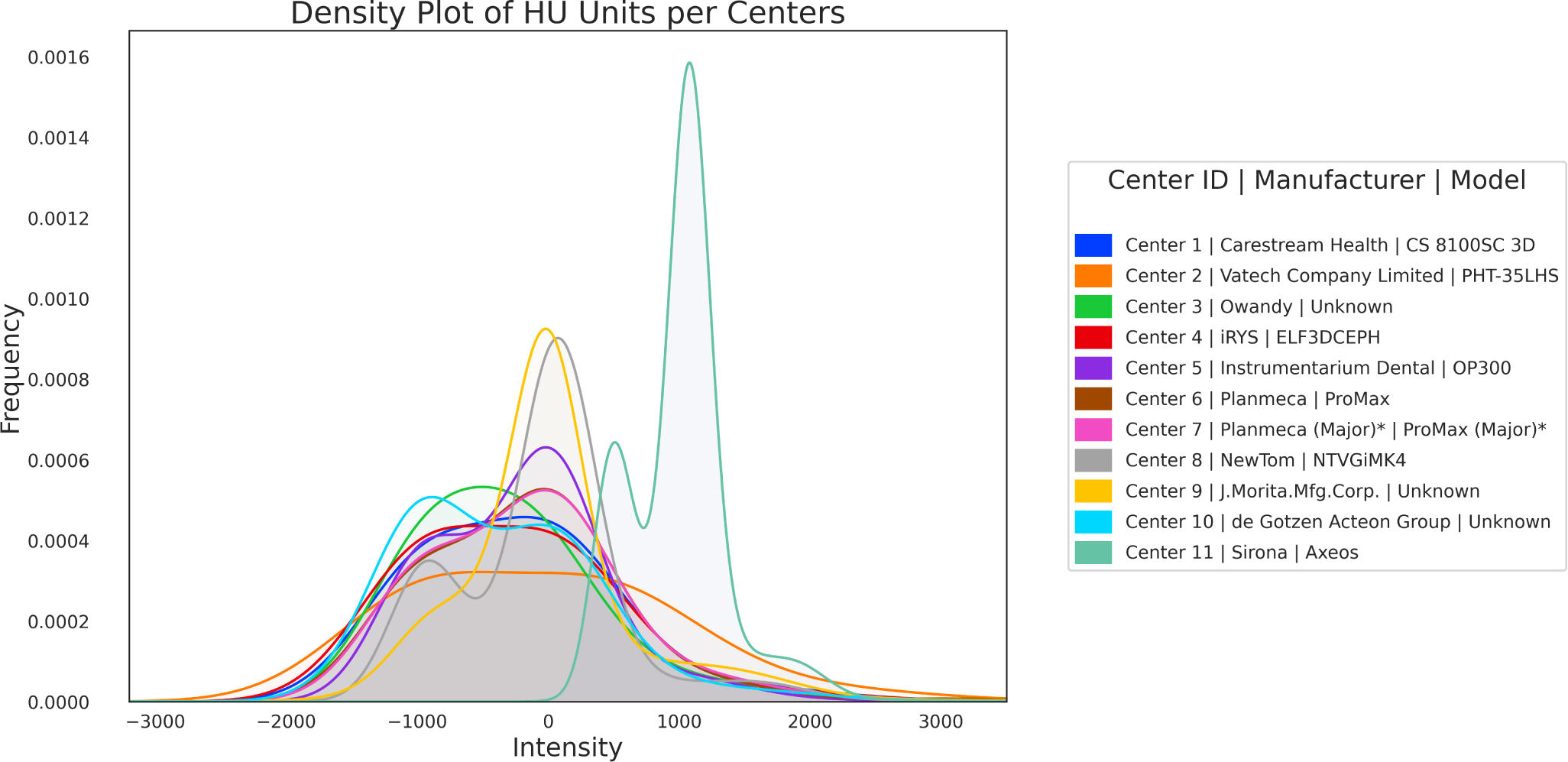
Distribution of intensity histograms across 11 centers in the study, illustrating variability in Hounsfield Units (HU). These variations reflect different radiation dosing protocols used by various manufacturers, emphasizing the importance of gathering a comprehensive dataset for creating a robust AI system. A total of 55 pairwise Kolmogorov-Smirnov tests were performed to evaluate the differences in intensity distributions between centers. P-values were adjusted for multiple comparisons using the Benjamini-Hochberg procedure. The asterisk (*) next to Center 7 indicates that the majority of its composition consists of Planmeca CBCTs, with some exceptions. For detailed information, refer to Table 1.

### Data Acquisition and Preparation

Developing deep learning applications for automated segmentation presents a significant challenge in ensuring model generalization across diverse dental practices and devices. To address this, we curated a comprehensive dataset of CBCT images from 11 different dental centers, covering a range of equipment from multiple manufacturers. The characteristics of the cohorts obtained from each dental center are shown in Table 1. Center 5 and Center 8 data derive from open-source datasets by Cui et al.^31^ and Cipriano et al.^19^, respectively, while the remaining centers represent French dental practices. In compliance with GDPR^32^, the dataset was anonymized in situ at each dental practice, rendering individual consent and ethical committee evaluation unnecessary by transforming the data beyond the scope of personal identification. Our study exclusively uses this deidentified dataset. Study cohorts are presented in detail in the Results section.

The variability in intensity densities, measured in Hounsfield Units (HU), across different centers is depicted in Figure 2. This highlights the inherent diversity in multi-centric data. Densities were estimated using histograms with 2000 bins for each center, and samples sizes varied, ranging from 9 to 156 (mean: 46). To assess the differences in intensity distribution across centers, pairwise comparisons were conducted using the Kolmogorov-Smirnov test. P-values were adjusted using the Benjamini-Hochberg procedure to control the increased risk of Type I error in the context of multiple comparisons.

### Ground Truth Annotation

Sinus and IAN regions, annotated on 358 and 499 CBCTs respectively, are detailed in Table 1. We employed Genesis^33^, a cloud-based annotation tool for medical imaging offering efficient 2D polygon interpolation. Annotations were conducted under guidelines developed with a senior dental practitioner with over 10 years of experience. The process began with 8 final-year dentistry students individually labeling each sinus and IAN voxels, with distinct classifications for left and right instances. Prior to the start of the annotation phase, the senior dentist provided training to each of the eight final-year dentistry students, aiming for consistent dataset annotations across the board. Following the initial annotations, experienced dental practitioners were tasked with reviewing and suggesting modifications. These suggestions were then implemented by the initial annotators to refine and ensure the accuracy of the dataset. Final validation of the data was dependent on approval from the senior dentist. The ground truth boundaries were obtained by applying a basic contour extraction method to the ground truth segmentation data.

### Data Pre-processing

In our experimental framework, we maintained consistent image resolutions across both stages for simplicity, although the architecture supports higher resolutions in the second stage. As a pre-processing step, volumes were all respaced to the 0.4mm resolution. Intensities were centered to exhibit a common mean value of −100, then clipped in the [−500, 2500] range and normalized in the [0, 1] range.

### Training details

#### Losses

Our model optimizes a loss function that combines five terms, effectively minimizing both segmentation, boundary and offsets regression errors. This function integrates a Dice Focal Loss^34^ for segmentation and boundary precision, an L1 loss for offset maps in the first stage, and similar error minimization for segmented and boundary-defined anatomy-centered cropped volumes in the second stage:

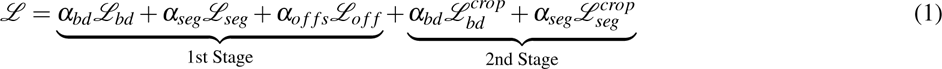

where

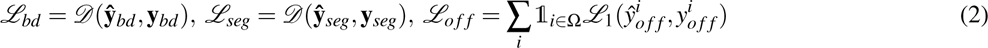

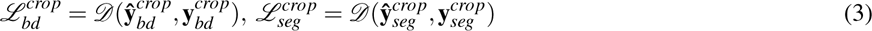

with

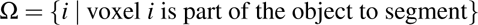

and

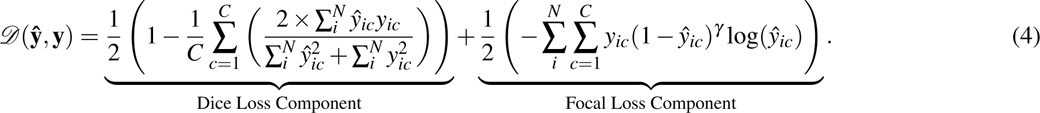

In this approach, 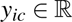 represents the true value for voxel *i* for class *c*, and 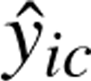 denotes the predicted probability for the same voxel. The focusing parameter is denoted by *γ*. *N* and *C* represent the total number of voxels and classes, respectively. The terms 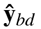 and **y***_bd_* correspond to the predicted and actual probability maps for boundary voxels. Similarly, 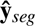 and **y***_seg_* refer to the predicted and actual probability maps for segmentation object voxels. The variables with the *crop* exponent refer to the tensors derived on the anatomy-centered crop volume. Finally, 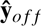 and **y***_o f f_* are the predicted and actual offset maps to the closest anatomy’s centroid along the *x*, *y*, *z* axes. Loss coefficients 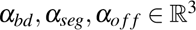 were selected to balance the weights between losses, with details provided in the Loss hyperparameters section of the Supplementary Materials.

#### Model implementation

Our models, implemented using Monai^35^ and PyTorch^36^ frameworks, were designed for automated sequential training of both stages. We used a computing setup featuring 32 GB RAM, an Intel Xeon Platinum 8259CL CPU (2.50GHz, 4 cores, 8 threads), and a Tesla T4 GPU with 15 GB memory. The first training stage, conducted over 200 epochs with a batch size of 3, was followed by a joint training of both stages (batch size of 1, 200 additional epochs). This sequential approach was necessitated by the dependency of stage two’s clustering algorithm on accurate initial stage predictions. A constant learning rate of 1e-4 was applied throughout the training. Stage one training took approximately 16 hours, and the joint training an additional 24 hours, cumulating in 40 hours total. During the training phase, the Crop Head processes randomly sized crops, selected from a predefined set and consistently centered on anatomical keypoints identified in the first stage. These crops vary from wide to narrow fields of view, targeting different anatomical areas. This approach is designed to improve the model’s robustness by providing exposure to a range of perspectives of anatomical regions. For inference, the model employs predetermined crop sizes to ensure complete encapsulation of the target objects: 64 x 64 x 64 mm^3^ for the sinus and 64 x 64 x 32 mm^3^ for the IAN. This strategy accounts for the standard orientation of CBCT images in the RAS (Right-Anterior-Superior) coordinate system. Further details on the Res-U-Net backbone, clustering algorithm hyperparameters, and training crop sizes are available in the Model architectures section of Supplementary Materials.

### Evaluation metrics

Volumetric and surface metrics were computed to evaluate segmentation accuracy in our study. For volumetric analysis, we used the Dice coefficient (5) and Intersection over Union (IoU) (5), which measure the overlap and similarity between segmented and ground truth volumes. For surface accuracy, we employed the Chamfer metric (6), assessing the average distance between predicted and actual contours, and the 95% Hausdorff Distance (HD95) metric (7), evaluating the maximum contour discrepancies.

The relevance of these metrics in model selection varied for different anatomical structures. For the sinus, a larger object, precise boundary delineation is key, making surface metrics like Chamfer and HD95 more pertinent. In contrast, the IAN, a smaller and more subjectively annotated structure, often exhibits discontinuities in annotation. In such case, volumetric metrics such as Dice and IoU give a better overall view, considering the possible variations in IAN segmentation.

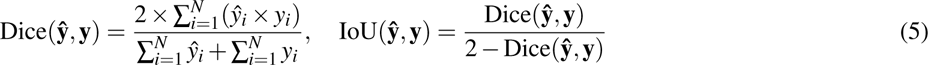

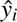 and *y_i_* are the predicted and ground truth values for the *i*-th voxel, respectively, and *N* is the total number of voxels.

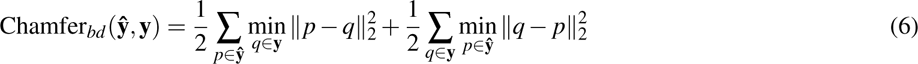

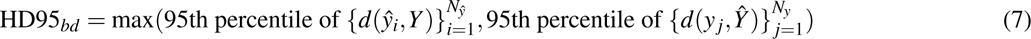

*d*(*x,Y*) denotes the minimum distance from point 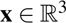 to any point in set 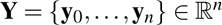, 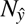_^_ and *N_y_* are the total number of points in the predicted and ground truth sets, respectively.

### Statistical analysis

In order to estimate the level of confidence in our results, 95% confidence intervals were computed using the bootstrapping method with 1000 replications. The Kolmogorov-Smirnov test was used to assess the variability in intensity distributions of CBCTs across different centers. The Benjamini-Hochberg procedure was used to adjust the K-S test for multiple comparisons. In our ablation study, Wilcoxon Signed-Rank test assessed performance differences among various models, ensuring each pairwise comparison was conducted using an identical dataset. The Mann-Whitney test was used in our generalization study to compare performance metrics on two independent datasets, the Internal Validation Set and the External Test Set. For comparing median annotation timings between experts and our SISTR model, the bootstrapping method with 1000 replications was used, appropriate for measures on two dependent and unpaired datasets. In this case, the p-value was derived from the distribution of mean differences across bootstrap samples. All tests were two-tailed and p-values < 0.005 were considered statistically significant. All p-values are reported in the results tables [2;3].

## Results

### Diversity in multi-centric data

An automated system using deep learning for outlining anatomical regions in radiology needs to be reliable and perform well across diverse cases. As shown in Figure 2, we analyzed the intensity distribution in Hounsfield Units (HU) across the various dental clinics and use 55 pairwise Kolmogorov-Smirnov tests in order to asses the variability of the centers. The statistical test were adjusted for multiple comparisons, based on Benjamini-Hochberg procedure and show statistical significance in HU distribution patterns among most dental centers (adjusted *p*-values < 0.005, see Table 2 in Supplementary Materials section for details). Notably, CBCT data from Sirona-manufatured machines exhibits distinct HU patterns compared to data from other manufacturers, as clearly evidence in Figure 2. This statistical evidence demonstrates the inherent heterogeneity in multi-centric CBCT data from 10 different dental CBCT manufacturers.

**Table 2.** Results of the Generalization Study with our SISTR method. This table summarizes the performance of the SISTR framework in segmenting the Sinus and Inferior Alveolar Nerve. Metrics are reported for both the Internal Validation Set and the External Test Set, further described in Table 1. A slight decrease in mean metric values is observed when comparing the Internal Validation Set to the External Test Set. However, the Mann-Whitney test reveals no statistically significant differences in metric distributions across these independent datasets. The term ‘ns’ denotes non-significant differences and is followed by the corresponding p-value in parentheses. The sample sizes for the External Test Set and the Internal Validation Set are also provided in parentheses, respectively.

| Dataset | DSC<sub>seg</sub> (%) | IoU<sub>seg</sub> (%) | DSC<sub>bound</sub> (%) | HD95<sub>bound</sub> (mm) | Chamfer<sub>bound</sub> (mm) |
| --- | --- | --- | --- | --- | --- |
| **Sinus** |  |  |  |  |  |
| Internal Validation Set | 97.41 (96.64-98.08) | 95.20 (93.97-96.43) | 80.34 (76.86-84.23) | 0.84 (0.60-1.17) | 0.23 (0.17-0.29) |
| External Test Set | 96.64 (95.38-97.60) | 94.36 (92.94-95.48) | 78.59 (76.16-81.03) | 1.62 (0.85-2.84) | 0.38 (0.24-0.60) |
| p-values | ns (0.92) (98, 31) | ns (0.92) (98, 31) | ns (0.54) (98, 31) | ns (0.79) (97, 31) | ns (0.67) (97, 31) |
| **Inferior Alveolar Nerve** |  |  |  |  |  |
| Internal Validation Set | 84.33 (81.64-86.66) | 77.39 (74.74-79.99) | 76.63 (74.06-78.84) | 5.45 (3.28-8.44) | 0.84 (0.50-1.37) |
| External Test Set | 83.43 (80.96-85.63) | 76.83 (74.56-79.16) | 75.98 (73.88-78.24) | 5.39 (3.62-7.53) | 0.88 (0.58-1.27) |
| p-values | ns (0.28) (91, 48) | ns (0.28) (91, 48) | ns (0.22) (91, 48) | ns (0.80) (83, 47) | ns (0.99) (83, 47) |

Table 1 reveals differences in key parameters such as Tube Voltage (kV) and Tube Current (mV) across the dental centers’ equipments. Notably, the input resolution of CBCTs, crucial in implantology and endodontics practices, varies significantly, with a range from 0.1 mm to 0.3 mm (median: 0.15 mm). Such variation underscores the need for high-precision segmentations finer than 0.4 mm, which single-stage methods struggle to achieve due to memory constraints when processing full-volume inputs. Our two-stage approach can effectively overcome this, enabling the second stage to achieve finer resolution through focused segmentation on a specifically cropped region of interest.

These variations in technical specifications and intensity distributions highlight the necessity of comprehensive and diverse datasets. Such datasets are essential for developing reliable AI models that aim at efficiency across a wide array of clinical settings and equipment types.

### Study cohorts

Detailed descriptions of the patient cohorts are provided in Table 1. We established two distinct study cohorts: the Ablation Study cohort and the Generalization Study cohort. For both cohorts, the data was divided as follows: 75% for the development set (of which 85% was used as the Internal Train Set and 15% as the Internal Validation Set), and the remaining 25% allocated to the Internal or External Test set, depending on the cohort.

In the Ablation Study cohort, both the development and testing phases involved data from the same centers. Conversely, the Generalization Study cohort used an External Test Set composed of centers that were completely independent of those in the development sets. For all experiments, model selection was based on the lowest loss during training as determined by the Internal Validation Set, with subsequent evaluations conducted on the appropriate Internal or External Test set.

### Segmentation performances

The segmentation performances of our SISTR method on the Ablation Study cohort are detailed in Table 3. Sinus segmentation achieved a strong precision, as evidenced by a 0.972 Dice score and a Chamfer boundary distance of 0.29 mm, which is lower than voxel precision of 0.4 mm. The IAN, a structure known for its challenging segmentation due to subjective annotation^25^, was segmented with a 0.855 Dice score and a Chamfer boundary distance of 0.63 mm.

**Table 3.** Experimental results of the Ablation Study. This table presents a comparison of segmentation and surface metrics for the Sinus and Inferior Alveolar Nerve segmentation task. Metrics are reported on the Internal Test Set of the Ablation Study cohort, while those obtained on the Internal Validation Set are available in Table 3 of the Supplementary Materials. A two-sided Wilcoxon Signed-Rank test was used to determine significant differences, with p-values detailed in subrows under each experiment. The baseline method for comparison is indicated in the respective column. Significance levels are marked with asterisks: *p < 0.005, **p < 0.001; The term ‘ns’ denotes non-significant differences and is followed by the corresponding p-value in parentheses. Sample sizes for the Internal Test Set are also provided in parentheses. Notably, the SISTR method demonstrates significant enhancements in segmentation tasks, particularly for the IAN dataset.

| Methods | Comparison | DSC<sub>seg</sub> (%) | IoU<sub>seg</sub> (%) | DSC<sub>bound</sub> (%) | HD95<sub>bound</sub> (mm) | Chamfer<sub>bound</sub> (mm) |
| --- | --- | --- | --- | --- | --- | --- |
| **Sinus** |  |  |  |  |  |  |
| base |  | 96.49 (95.91-97.01) | 94.49 (93.84-95.12) | 76.22 (74.28-78.23) | 1.82 (1.39-2.40) | 0.39 (0.33-0.46) |
| base<sub>bd</sub> |  | 96.54 (96.05-97.01) | 93.66 (92.88-94.49) | 76.04 (74.04-78.15) | 1.98 (1.49-2.71) | 0.40 (0.35-0.45) |
|  | vs base | ns (0.35) (82) | ns (0.30) (82) | ns (0.27) (82) | ns (0.02) (75) | ns (0.70) (75) |
| sistr (ours) |  | 97.17 (96.56-97.63) | 94.79 (93.91-95.60) | 78.02 (75.86-80.16) | 1.06 (0.81-1.37) | 0.29 (0.23-0.35) |
|  | vs base | < 0.001** (82) | < 0.001** (82) | < 0.001** (77) | < 0.001** (77) | < 0.001** (77) |
| sistr<sub>bd</sub> |  | 97.18 (96.61-97.60) | 94.79 (93.98-95.58) | 79.47 (77.63-81.30) | 1.14 (0.86-1.46) | 0.27 (0.22-0.32) |
|  | vs sistr | ns (0.47) (82) | ns (0.44) (82) | < 0.001** (82) | ns (0.56)(82) | < 0.005* (82) |
| sistr<sub>da</sub> |  | 97.16 (96.66-97.58) | 94.73 (93.94-95.50) | 77.68 (75.61-79.69) | 1.13 (0.88-1.42) | 0.30 (0.25-0.35) |
|  | vs sistr | ns (0.05) (82) | ns (0.19) (82) | ns (0.22) (82) | < 0.001** (81) | < 0.001** (81) |
| **Inferior Alveolar Nerve** |  |  |  |  |  |  |
| base |  | 75.17 (73.41-77.06) | 68.29 (66.17-70.48) | 68.11 (66.10-70.65) | 6.078 (4.767-7.423) | 1.16 (1.04-1.29) |
| base<sub>bd</sub> |  | 74.47 (72.49-76.47) | 67.82 (65.56-70.08) | 68.29 (66.19-70.93) | 4.650 (3.983-5.513) | 0.97 (0.89-1.06) |
|  | vs base | ns (0.07) (96) | ns (0.09) (96) | ns (0.43) (96) | ns (0.07) (89) | < 0.001** (89) |
| sistr (ours) |  | 85.50 (83.78-87.05) | 78.65 (76.78-80.49) | 78.22 (76.40-79.94) | 4.929 (3.890-5.976) | 0.63 (0.50-0.77) |
|  | vs base | < 0.001** (96) | 0.001** (96) | < 0.001** (96) | ns (0.14) (89) | < 0.001** (77) |
| sistr<sub>bd</sub> |  | 84.34 (82.71-85.99) | 77.32 (75.33-79.30) | 78.08 (76.28-79.86) | 6.468 (5.035- 7.970) | 0.90 (0.65-1.21) |
|  | vs sistr | ns (0.01) (96) | ns (0.01) (96) | ns (0.67) (96) | ns (0.04) (94) | ns (0.06) (96) |
| sistr<sub>da</sub> |  | 84.12 (82.33-85.94) | 77.19 (75.34-79.22) | 76.88 (74.91-78.75) | 6.82 (5.21-8.84) | 0.96 (0.67-1.35) |
|  | vs sistr | < 0.001** (96) | < 0.001** (96) | < 0.001** (96) | ns (0.03) (93) | < 0.005** (93) |

Figure 3 displays the model’s performance over several clinical scenarios, from simple to more challenging cases, such as those involving artifacts or edentulous arches. For practical clinical application, it is imperative that our automated segmentation tool not only performs well in common situations but also effectively adapts to less frequent yet real-world occurrences. From Figure 3, it can be seen that the model is able to accurately segment the sinuses in cases with maxillary artefacts and to provide accurate, uninterrupted delineation of the IAN in cases of missing teeth in the mandible and in edentulous arches. These test set examples, derived from CBCTs scans sourced from various manufacturers (Morita, Planmeca, Carestream, Vatech), highlight the model’s robustness and its capacity for generalization across diverse clinical conditions.

**Figure 3.**
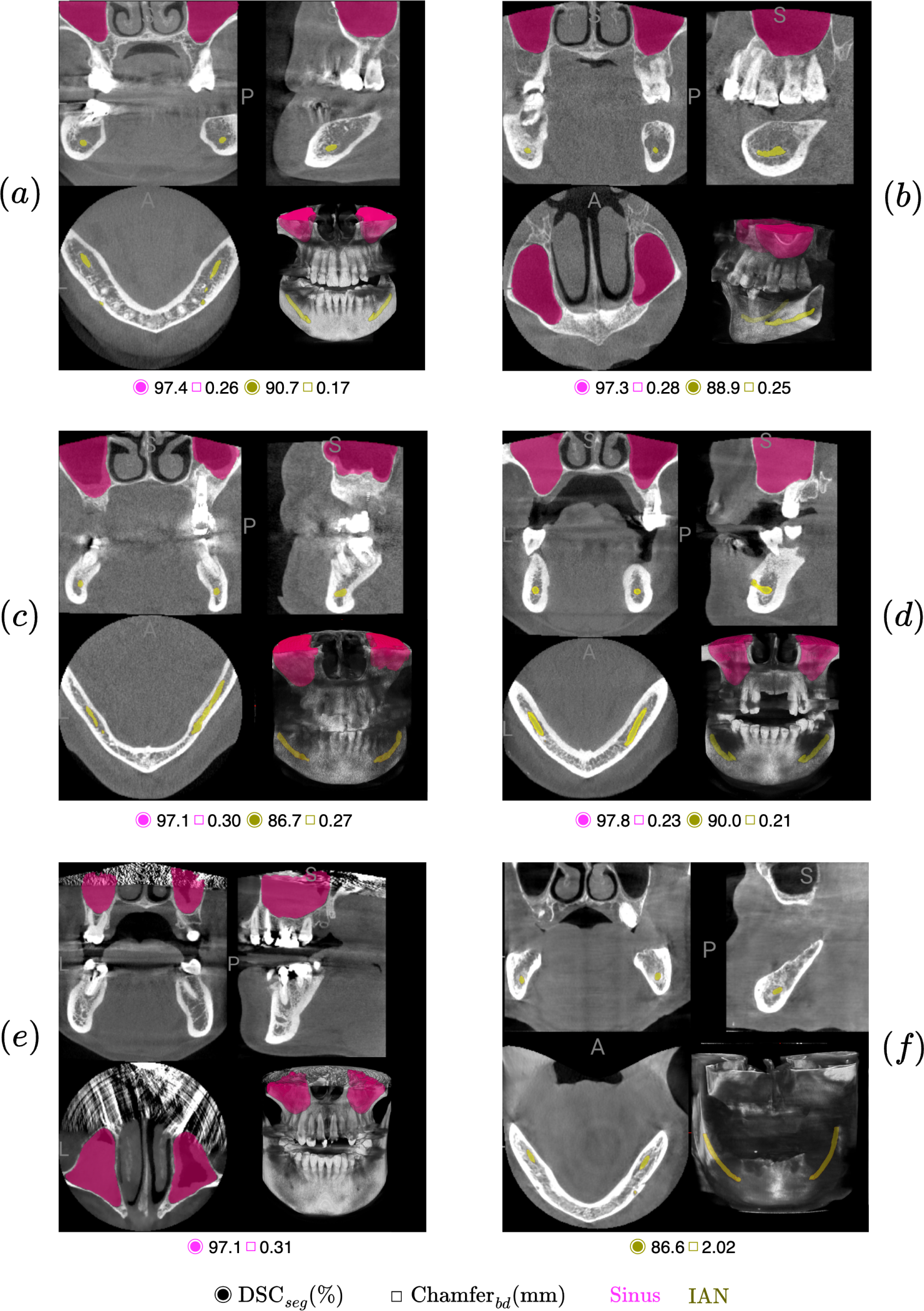
Evaluation of our SISTR method in various challenging scenarios. This figure demonstrates the performance of our SISTR model on diverse cases from the Internal Test Set of the Ablation Study cohort. For each case, the Dice score for segmentation (DSC_seg_(%)) and Chamfer distance for boundary (Chamfer_bd_(*mm*)) are provided. The cases include: (a) a Morita scan with artefacts; (b) a Planmeca scan of an arch with missing teeth; (c) a Planmeca scan with severe artefacts due to implants; (d) a high-quality Carestream scan; (e) a Carestream scan with significant artefacts in the maxillary region; (f) a Vatech scan of edentulous arches. Notably, cases (a) to (d) are included in both the sinus and IAN test sets, while cases (e) and (f) are exclusive to the sinus and IAN test sets, respectively. Notably, our model demonstrates robust performance across most cases, effectively handling even the more challenging scenarios.

### Generalization study

To evaluate the generalization capabilities of our method, we conducted experiments using the Generalization Study cohort, where the development and test sets feature entirely independent centers. Models were trained on CBCTs sourced from 7 centers and externally tested on CBCTs sourced from 4 unseen centers. The performance results of our SISTR method on both the Internal Validation Set and the External Test Set are detailed in Table 2. These results indicate the model’s ability to generalize effectively to unseen data. This is evidenced by a modest decrease of one percentage point in the Dice score performance, declining from 0.974 to 0.966 for the Sinus segmentation task, and from 0.843 to 0.834 for the IAN segmentation task. Furthermore, the Mann-Whitney U test, applied to assess performance differences on the independent test set, did not reveal any statistical differences in the distribution of metrics between the two evaluation sets. This finding reinforces the potential of our SISTR method for broad applicability in diverse real-world scenarios.

### Ablation studies

We conducted an extensive ablation study focusing on the novel components of our framework. This study involved the integration of a boundary prediction branch and our distinctive dual-stage approach. Models were developed and evaluated using the Ablation Study cohort described in Table 1.

#### Description

For baseline comparison, we developed a straightforward, single-stage model named *base*. This model directly segments resized and padded volumes ensuring a uniform input size. The objective of this baseline model is to understand the extent of improvements imputable to the localization module (stage 1) of our SISTR method by using complete input volumes without specific regions of interest. Our second ablation study introduces a boundary prediction branch to the *base* model, resulting in the *base_bd_* variant, to assess the impact of the boundary feature on segmentation accuracy. Both the baseline single-stage models underwent training for 200 epochs, mirroring the duration of the first stage training in the *sistr* approach. The third experiment, *sistr_bd_*, incorporates the boundary prediction into SISTR’s second stage, aiming to refine segmentation output. The final experiment, *sistr_da_* adds extensive data augmentation techniques— in SISTR’s second stage such as random flips, rotations, coarse dropout, and Gaussian and Gibbs noise—in the second stage to explore potential enhancements in model robustness.

The reported metrics were computed using checkpoints for which the segmentation loss was minimized on the Internal Validation Set, *L_seg_* for single-stage models and 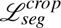 for dual-stage models, respectively.

#### Metrics computation details

For experiments *base_bd_* and *sistr_bd_*, the reported results for all DSC*_bound_*, HD95*_bound_* and Chamfer*_bound_* metrics are computed on the boundaries predicted by the model. Whereas for experiments *base* and *sistr*, those metrics are computed on boundaries extracted from the predicted segmentation. To make the evaluation consistent and comparable, all reported metrics were computed on segmentation results standardized to a volume resolution of 0.4 x 0.4 x 0.4 mm^3^ per voxel. This means for approaches *sistr* and *sistr_bd_*, both the left and right maxillary sinus (resp. IAN) were predicted on the organ-centered refined volume (second stage) and translated back to the native volume coordinates. This conversion enables us to report metrics in mm, facilitating comparison between approaches. To evaluate significant differences between experiments, a two-sided Wilcoxon Signed Rank test was conducted comparing each experiment with its respective baseline (namely, *base_bd_* vs. *base*, *sistr* vs. *base*, *sistr_bd_* vs. *sistr*, and *sistr_da_* vs. *sistr*). The p-values are reported in Table 3 with a threshold of p < 0.005 to denote significant differences in metric medians, thus indicating statistically significant variations among the experiments.

#### Results

The outcomes of this ablation study are summarized in Table 3. We illustrate the statistics of our ablation study in Figure 4 displaying examples of respectively high, medium, and low-performing predictions. Additionally, Figure 5 and Figure 6 illustrate segmentation predictions of the different models used in the ablation study.

**Figure 4.**
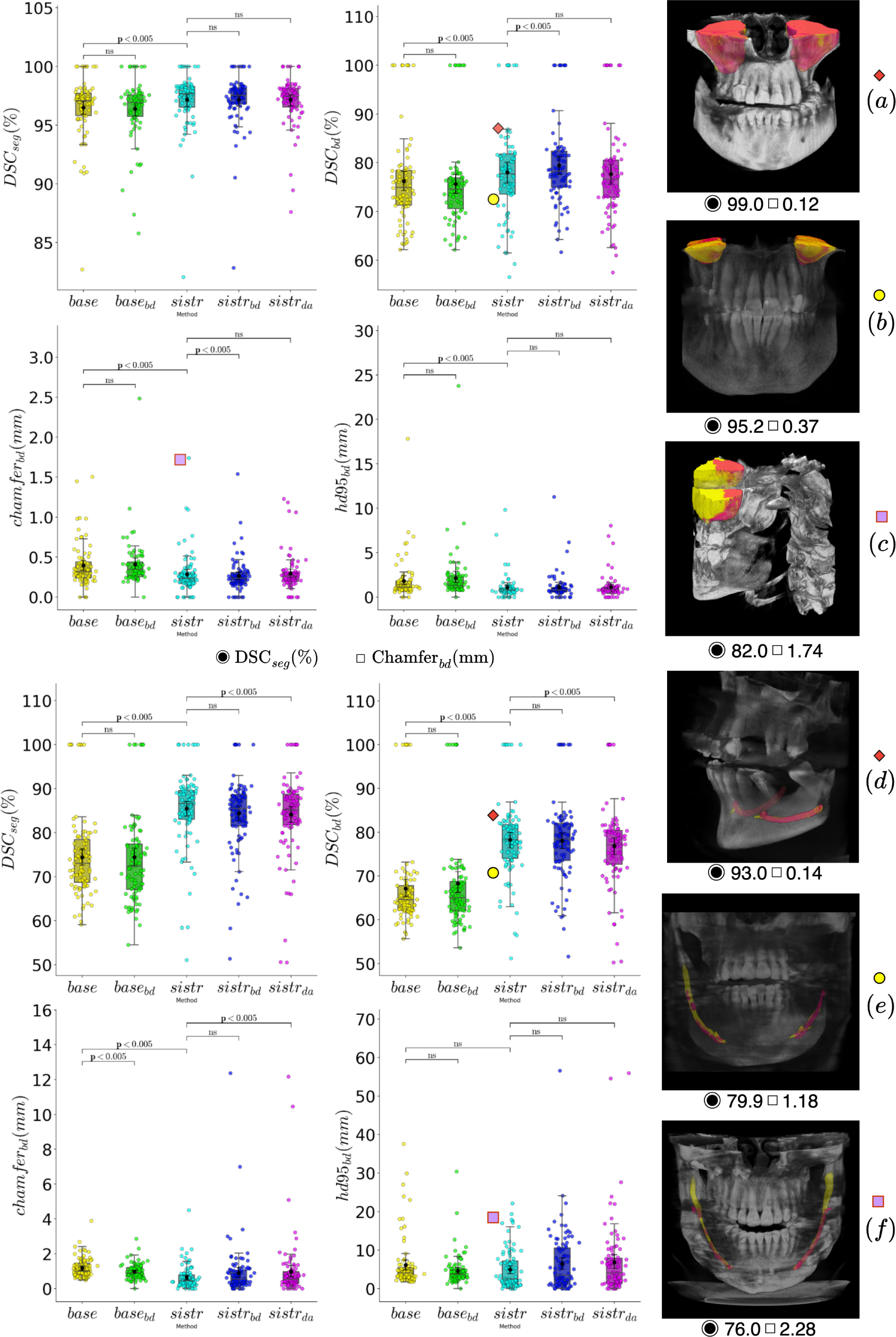
Quantitative ablation studies and qualitative evaluation for maxillary sinus and IAN segmentation using the SISTR method. (Top): Boxplot detailing performances of each model in the ablation study for maxillary sinus segmentation, on four specific metrics: DSC*_seg_*(%), DSC*_bd_* (%), Chamfer*_bd_* (*mm*) and HD95*_bd_* (*mm*). Panels (*a−c*) present SISTR’s segmentation results, varying in accuracy, with a strong volumetric Dice score (*a*), a medium boundary Dice score (*b*) and a poor boundary Chamfer (*c*). (Bottom): Similar boxplot for IAN segmentation. Panels (*d − f*) display outcomes with different Dice scores and HD95 metrics. For all visual examples, ground truth is indicated in yellow; SISTR predictions are in pink-red. The boxplots highlight statistically significant differences between models, indicated by *p*-values from Wilcoxon Signed-Rank test; the term ‘ns’ denotes non-significant differences. Boxes are representing the inter quartile range (IQR), extending from Q1 to Q3 and centered on the median value. Upper whiskers represent the highest data point that is less than Q3+1.5 IQR. Lower whiskers represent the smallest data point that is greater than Q11.5 IQR. Panel (*c*) features a partially cropped CBCT, potentially explaining the low prediction accuracy. Panel (*f*) highlights the model’s omission of the IAN’s distal part, an area prone to annotation variability among experts^14^.

**Figure 5.**
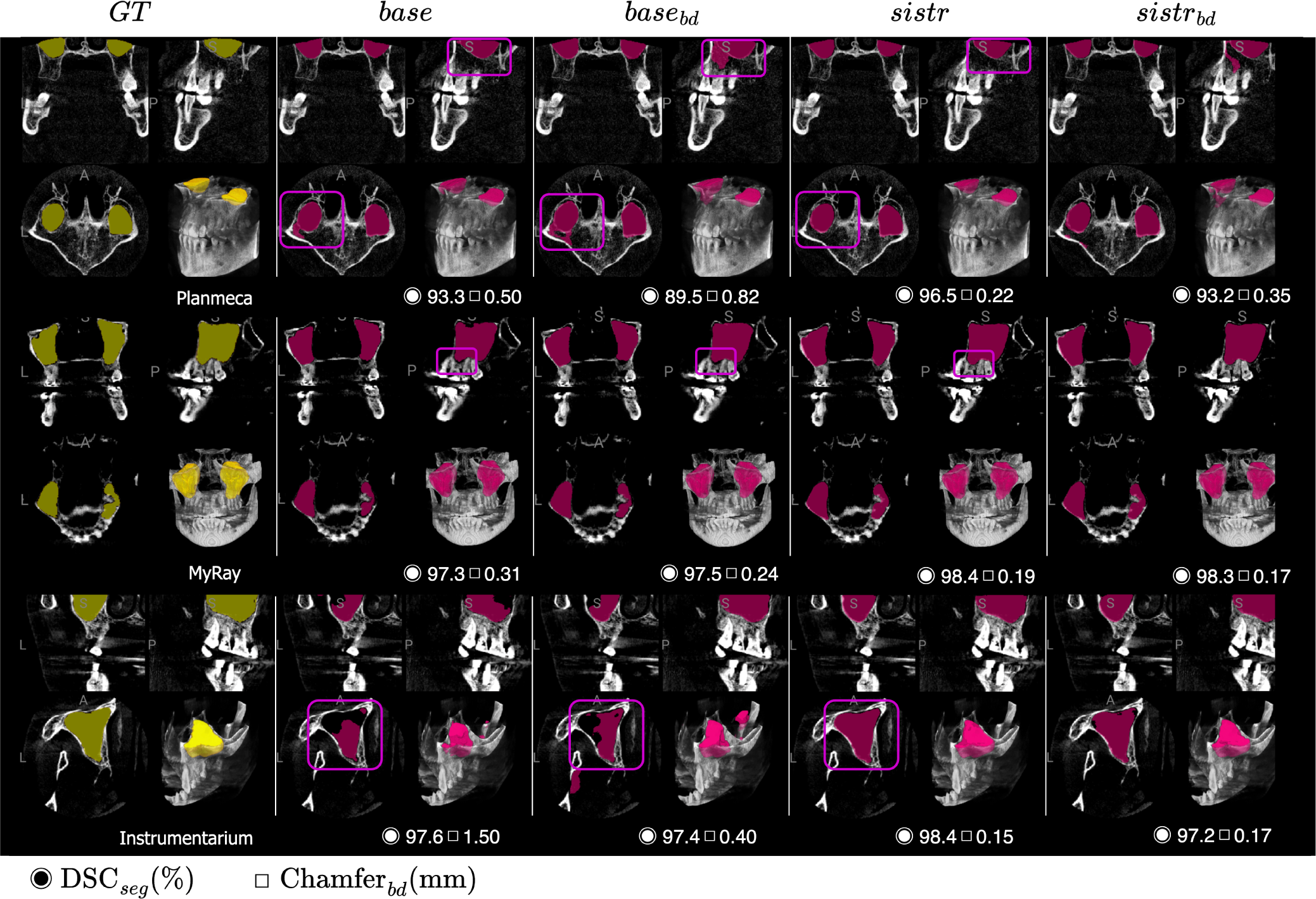
Qualitative segmentation results of maxillary sinus for the comparative models on various providers. Visual examples of the different models in our ablation studies are shown. Ground truth is in yellow; models prediction in pink-red. Performance metrics are reported underneath each examples while manufacturer brands are reported underneath each ground truth image. Our model, *sistr*, significantly improves sinus delineation on the Instrumentarium truncated CBCT showcased in the third row. It also reduces false positive prediction on the Planmeca CBCT as shown in the first row. Overall, *base* predictions are quite accurate, as already suggested by metrics in Table 3, making the small improvements brought by *sistr* hard to notice.

**Figure 6.**
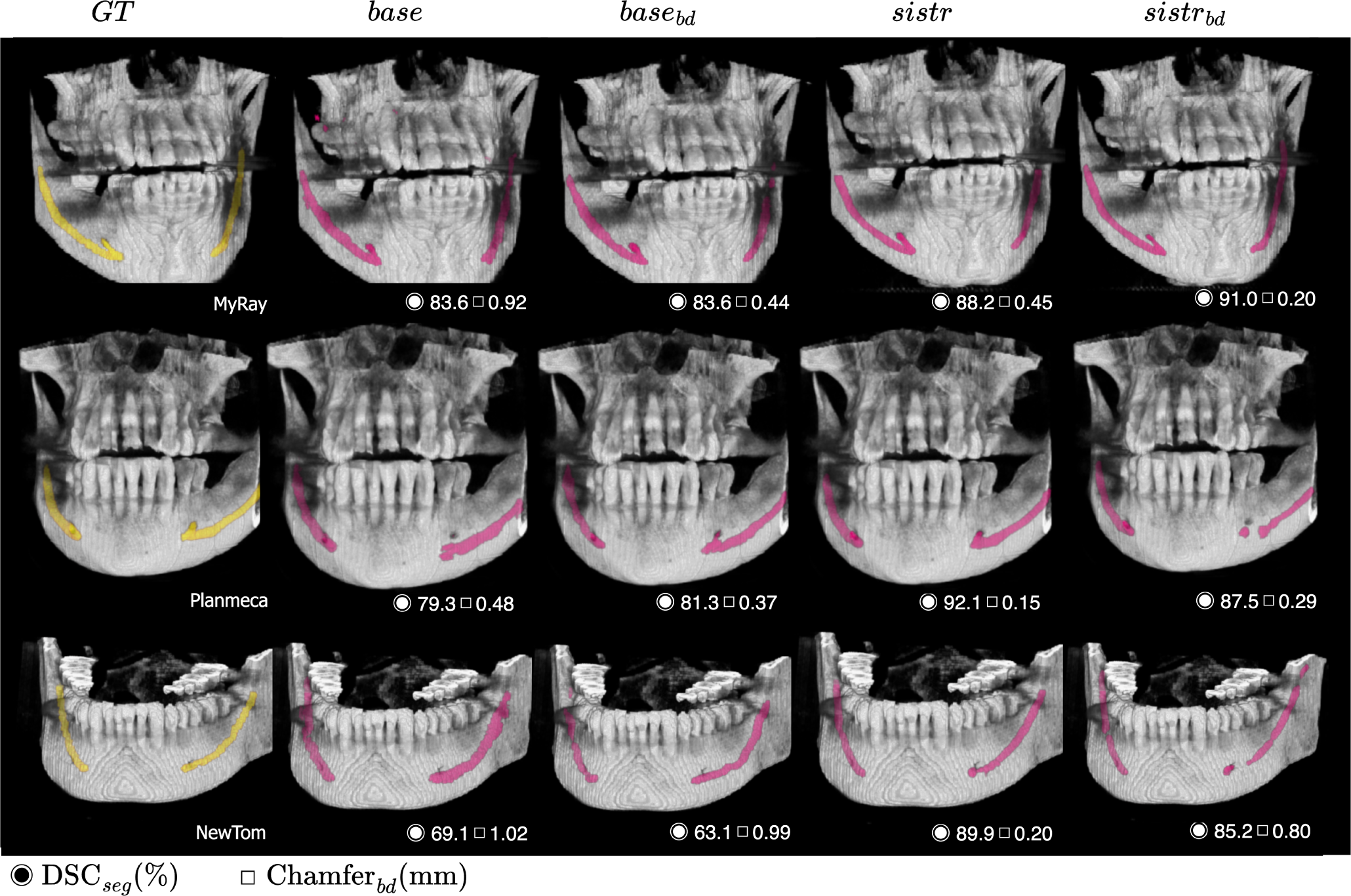
Qualitative segmentation results of IAN for the comparative models on various providers. Visual examples of the different models in our ablation studies are shown. Ground truth is in yellow; models prediction in pink-red. Performance metrics are reported underneath each examples while manufacturer brands are reported underneath each ground truth image. One can notice false positive prediction on the Myray example (first row) for the *base* prediction, while *sistr* significantly improves the prediction especially in the right distal quadrant. Visually, *base* predictions are quite coarse and oversegmented in comparison to those from *sistr* (Planmeca, NewTom). Detailed segmentation of the mental foramen is achieved by *sistr*, while *base* misses this region (Planmeca, second row).

Our method significantly improves both volumetric and surface metrics for IAN and sinus segmentation tasks compared to the *base* and *base_bd_* models. For the IAN segmentation, it demonstrates a notable 10% increase in DSC_seg_ (from 0.750 to 0.855) and a 45% reduction (0.5 mm) in the Chamfer metric (p < 0.005). In sinus segmentation, while the increase in segmentation Dice Score is modest (0.007 increment in DSC_seg_ from an already high baseline of 0.965), there is a significant improvement in DSC_bd_ by 0.02 points and a 33% reduction (0.1 mm) in the Chamfer metric (p < 0.005 for all metrics). These results suggest that even with minor volumetric gains, especially in large volumes like the sinus, our method achieves significant boundary improvements, as evidenced by the Chamfer metric. This is further illustrated in Figure 5 (third row; Instrumentarium case), highlighting the method’s effectiveness in enhancing segmentation accuracy.

Incorporating a boundary prediction branch into the segmentation head, either coarse for the single-stage or focused on anatomy-centered cropped zones for the dual-stage model, did not result in substantial improvements in surface or volume metrics. This observation is corroborated by statistical analyses comparing *base_bd_*with *base* and *sistr_bd_* with *sistr*, which revealed high p-values, indicating no statistically significant differences. Yet, it is important to highlight that *base_bd_* exhibited a notable improvement in boundary Chamfer metrics, displaying a reduction of 0.2mm in Chamfer*_bd_* (*p*-value < 0.005) compared to the baseline *base*. However, the inclusion of data augmentation in *sistr_da_* did not yield the anticipated improvements, as indicated by the minimal changes observed in performance metrics. As visually demonstrated in Figure 6, our approach significantly enhances the continuity of segmented structures compared to a basic baseline, a crucial aspect for accuracy and safety in implant planning.

### Clinical endpoints

To introduce preliminary elements towards the clinical validation of our novel segmentation methodology for the Sinus and the IAN, we conducted a study with a focus on two initial endpoints:

1. Comparison of the time efficiency between semi-automatic segmentation with manual intervention and fully automated AI-driven segmentation.
2. Analysis of the inter-observer variability among experts in segmenting the IAN.

For the first endpoint, we gathered data logs from our annotation platform used by nine annotators tasked with segmenting the Sinus and IAN. The volume of cases segmented by each annotator ranged from 7 to 43 for the Sinus and 10 to 37 for the IAN. Concurrently, we monitored the inference time of our AI-based SISTR method on datasets comprising 82 and 96 CBCT scans for Sinus and IAN segmentation, respectively. On average, semi-automatic manual segmentation required 33.7 minutes (range: 30.2-37.1 minutes) for the Sinus and 25.8 minutes (range: 22.9-29.2 minutes) for the IAN, with standard deviations of 10.7 and 9.4 minutes, respectively. The fastest annotator completed these tasks in 20.8 minutes for the Sinus and 14.2 minutes for the IAN. In contrast, the SISTR AI models displayed expected efficiency, with average inference times of just 0.071 minutes (4.36 seconds) and 0.065 minutes (3.9 seconds) for the Sinus and IAN, respectively. This includes all stages such as model and data loading, preprocessing, and inference. A bootstrapping statistical test, employing 1000 replications, confirmed a significant difference in processing times between the fastest human annotator and the AI model (p-value = 0.0 for both anatomical regions). These results, while anticipated, effectively illustrate the advantage of AI in terms of time efficiency compared to non-automated methods. All these results are shown on Figure 7.

**Figure 7.**
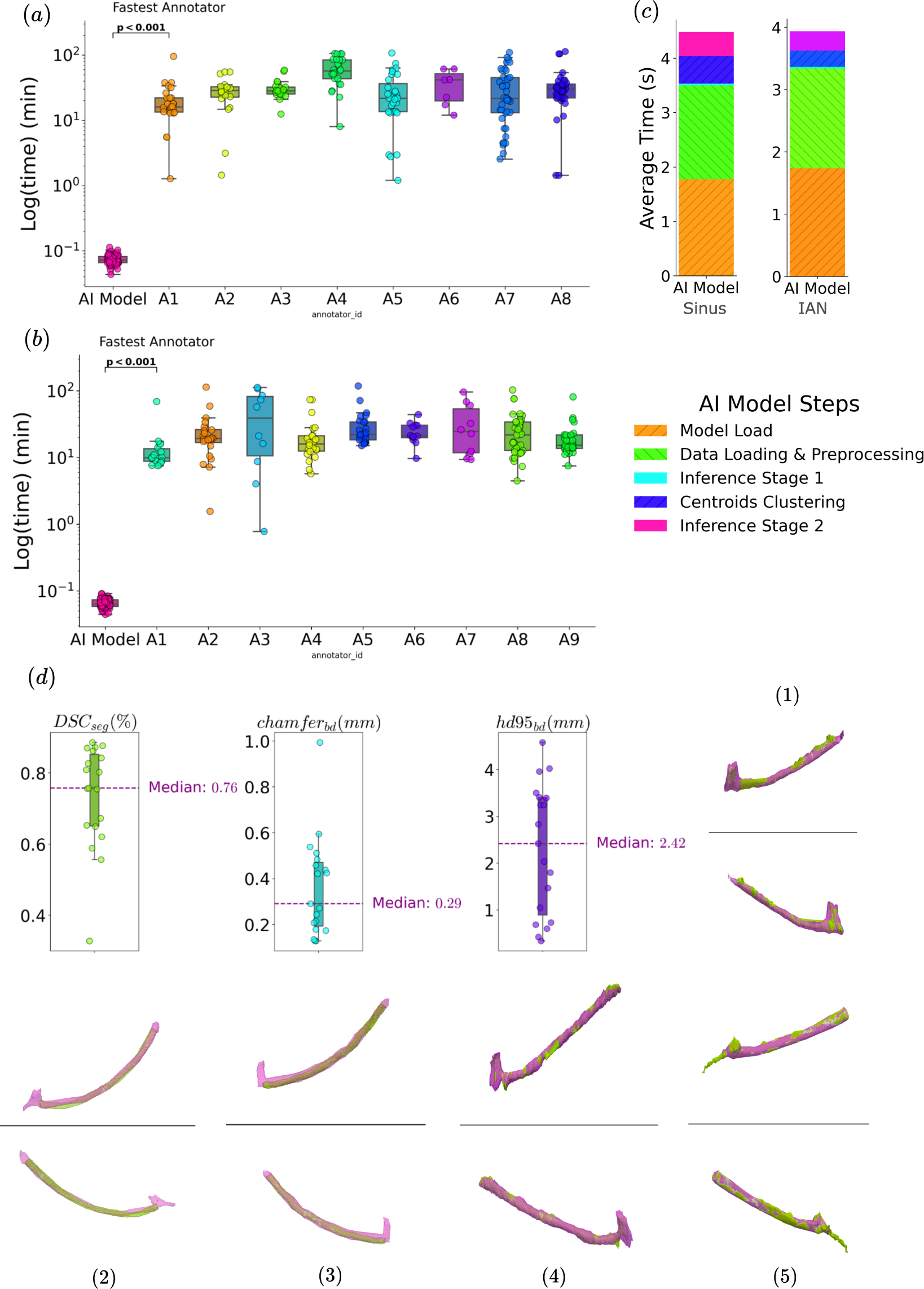
Comparison of time efficiency between our AI-based SISTR segmentation and semi-automatic manual segmentation, and analysis of inter-observer variability in IAN Segmentation. (a) and (b) display the processing times for each annotator (A1 to A9) and our AI model for Sinus and IAN segmentation, respectively. The number of cases annotated by each of the eight or nine annotators ranged from 7 to 45 per anatomical region. Bootstrapping analysis revealed a significant difference in processing times between the fastest annotator (A1) and the AI model (p-value = 0.0). The average manual segmentation time was 33.7 minutes for the Sinus and 25.8 minutes for the IAN, compared to the SISTR AI Model’s average of 0.071 and 0.065 minutes, assessed on datasets of 82 and 96 samples, respectively. (c) details the processing times for all the stages of our AI model, including model and data loading, preprocessing, and inference, with total inference times averaging approximately 4 seconds. (d) examines the inter-observer variability in IAN segmentation, based on 7 CBCTs and 18 pairs of observations, yielding a mean Dice score of 0.758 (range: 0.709-0.804) and a mean Chamfer distance of 0.29 (range: 0.252-0.396). Superimpositions (in pink and light green) of identical case segmentations by different experts, shown in (1) to (5), demonstrate increased variability and disagreement primarily in the mandibular foramen region and the distal end of the IAN.

Regarding the second endpoint, the inter-observer variability for IAN segmentation was examined using 7 CBCTs, annotated by different experts, yielding 18 pairs of comparative analyses. The mean Dice coefficient for these comparisons was 0.758 (range: 0.709-0.804), whereas our method reached an average Dice score of 0.834 (range: 0.810-0.856) on an external test set. Additionally, we identified the mandibular foramen region and the distal end of the IAN as areas particularly prone to annotation subjectivity, as evidenced in illustrative examples in Figure 7, captions (1) to (5).

These results not only highlight the satisfactory performance of our model, particularly in the context of inherent annotation variability, but also its efficiency in processing time.

## Discussion

We have developed a unified framework for the precise segmentation of the maxillary sinus and the inferior alveolar nerve (IAN) from Cone-Beam Computed Tomography (CBCT) images, leveraging a dataset representative of 11 dental clinics. This framework, externally validated on an independent test set comprising 98 and 91 CBCT scans for the sinus and IAN respectively, is inspired by the work of Cui et al.^26^, initially designed for teeth segmentation. Our methodology has been found to be effective in dealing with anatomical regions that have multiple instances in the oral cavity, such as the sinus, IAN, and teeth. This is achieved through instance localization using the centroids offsets regression module of SISTR.

SISTR has demonstrated greater accuracy than single-stage methods for both anatomical regions. The study confirms that focusing segmentation efforts on specific regions of interest leads to improved results. This improvement is primarily due to two factors: the reduction of false positives and the enhancement of the model’s adaptability to specific subvolumes. These targeted subvolumes typically exhibit less variability than entire CBCT volumes, which often have varying Field-Of-Views. Our work excels in IAN segmentation, as SISTR achieves a significant increase of over 10 percentage points in Dice Score compared to the single-stage baseline. The slight improvement in sinus segmentation, indicated by a 0.5 percentage point increase in Dice Score, may be attributed to the relatively simpler nature of the task compared to others, as suggested by the baseline models’ results. The literature acknowledges the complexity of IAN segmentation, largely due to considerable inter-observer and intra-observer variability; for instance, Järnstedt et al.^14^ reported an inter-observer variability of 0.77 mm, benchmarked against a gold standard set by experienced radiologists. Comparative studies like Cipriano et al.’s^25^ and Usman et al.’s^24^ used internal test sets with limited diversity, primarily from one or two centers, often overlapping with their development sets. This lack of external validation and diversity in test datasets limits the generalizability of their findings. For instance, Cipriano et al.^25^ achieved a Dice score of 0.79 using a multi-tier training approach. This was based on 256 synthetic annotations, 68 actual annotations, and an internal test set of 15 CBCTs, although limited by the use of a single data source. In contrast, Usman et al.^24^ reported Dice scores of 0.751 and 0.77 on an internal test set of 500 CBCTs from a unique center and on Cipriano et al.’s dataset^25^, respectively, employing a development set of 500 densely annotated CBCTs from a single center. While these results are noteworthy, they are in a different range of magnitude compared to our achieved Dice score of 0.843 (range: 0.810-0.856), underscoring the potential superiority of our method. A definitive comparison would require identical development and test sets.

Regarding Sinus segmentation, Morgan et al.^17^ developed an automatic model achieving an impressive Dice score of 0.984, tested internally on 30 scans from two centers. Their results surpass ours in terms of magnitude, but are constrained by a smaller dataset and reduced variability, with scans originating from only two manufacturers.

Our study prioritizes data diversity over sheer volume, which distinguishes it from similar published works. While this focus may not be commonly observed in similar research, we believe it is an important aspect to consider. We compiled a comprehensive dataset from 11 dental clinics, featuring 358 and 499 CBCT scans for sinus and IAN, respectively, from both seen and unseen centers. This approach contrasts with the work of Cui et al.^26^, which, although vast (4938 CBCTs from fifteen centers), does not clearly demonstrate the necessity of such volume for accuracy. Our emphasis on varied data sources rather than quantity aims to enhance the generalizability and robustness of our segmentation method. Additionally, our model significantly outpaces semi-automatic segmentation methods, delivering results in an average of 0.06 minutes (3.6 seconds) compared to the 25 to 30 minutes required for manual segmentation, underscoring the efficiency of AI-based systems.

Our approach has shown significant results in distinguishing the maxillary sinus and IAN from CBCT images. However, its effectiveness may be limited in certain cases due to its dependence on specific anatomical features. This limitation is particularly evident in the model’s need for tailored calibration during the clustering phase, as it must be adjusted for different anatomical structures. The clustering algorithm used in our model to identify centroids is sensitive to anatomical variations, which requires customized parameter settings for each structure. To address this limitation, we propose incorporating a Multi-Layer Perceptron (MLP) that would enable the direct derivation of centroids from the predicted offset map, potentially enhancing the model’s adaptability. The current methodology requires manual parameter selection for anatomical object cropping, which limits its generalizability as a framework. A promising refinement here could be the introduction of an inductive bias. By employing the coarse segmentation prediction from the initial stage to inform object cropping, the model could achieve a higher degree of automation and precision.

Considering future advancements, reevaluating the current centroid-based phase and exploring the integration of skeleton information could yield substantial improvements. The innovative approach by Cui et al.^26^, which employs morphological features like centroids and skeletons for tooth segmentation, offers an insightful reference. Adapting similar techniques for IAN segmentation could lead to enhanced object localization in the initial stage, coupled with more precise delineation, especially at the extremities.

Another aspect of our method that could be improved is the resolution in the latter stage of the two-stage process. Although the first stage efficiently performs coarse segmentation at lower resolutions, increasing the resolution in the second stage could significantly refine the segmentation quality, allowing for more detailed and intricate delineations.

Transforming the two-stage training process into an end-to-end workflow could yield benefits in metrics, development time, and inference time. Currently, the initial stage of SISTR disrupts the differentiability of the pipeline by involving cropping of the original volume around predicted centroids. To achieve a seamless end-to-end structure, shifting towards soft attention, instead of hard attention, could be beneficial, though it might result in some resolution loss. The potential solution worth exploring in this context is the use of an Attention-Unet architecture^37^.

Expanding our model to include multi-class segmentation is another way to improve it. By adding bottlenecks that capture positional information within the entire image^25,38^, we can transform our instance segmentation model into a more comprehensive multi-class segmentation framework.

Although our study provides a strong foundation for evaluating the clinical relevance of our segmentation methodology, it is important to note that this is only the first step in a comprehensive clinical assessment. To gain a more complete understanding of the effectiveness of our method, future research should prioritize comparing the accuracy of segmentations and detections made solely by human experts against those augmented by AI, with this comparison as a key endpoint. A more extensive examination involving a larger number of CBCTs, annotated by multiple experts, would provide more profound insights on inter-observer variability in IAN segmentation, thus enhancing the clinical applicability of our method.

## Author contributions statement

Study methodology, conception and design: L.M, E.C and H.M. Software: L.M, E.C., H.M. and W.A. Data collection: L.M. and H.M. Analysis and interpretation of results: L.M., E.C, H.M., W.A., J.R., C.A. Draft manuscript preparation: L.M., E.C, H.M. Manuscript review: L.M., E.C., H.M., W.A., J.R., C.A. Supervision: L.M. All authors were involved in critical revisions of the manuscript, and have read and approved the final version.

## Data availability

The full datasets are protected because of privacy issues and regulation policies in dental clinics. However part of the data is available in the following public domain resources: https://pan.baidu.com/s/1LdyUA2QZvmU6ncXKl_bDTw, and https://ditto.ing.unimore.it/maxillo/dataset/

## Code availability

Code is fully available through the corresponding author upon reasonable request.

