## Supplementary Materials for "SISTR: Sinus and Inferior alveolar nerve Segmentation with Targeted Refinement on Cone Beam Computed Tomography images"

### Centroid clustering hyperparameters

| Model | Minimum votes per voxel | Minimum voxel distance between peakpoints | Minimum weight score |
|---|---|---|---|
| IAN | 20 | 70 | 20 |
| Sinus | 25 | 10 | 25 |

**Table 1. Centroid clustering hyperparameters:** this table presents the clustering parameters of our SISTR method for the Sinus and Inferior Alveolar Nerve models.

The clustering algorithm, known as the Fast Search algorithm, introduced by Rodriguez, A. & Laio, A., 2014, uses several hyperparameters to determine candidate centroids. A voting map gathers the number of times given voxels were designated by their neighboring voxels as the target centroid. Below is a quick explanation of each hyperparameter:

- **Minimum votes per voxel**: The minimum number of votes that each voxel must receive to be considered as a potential centroid. This threshold serves as a first filter on the candidate centroids, based on the voting map.
- **Minimal voxel distance between peak points**: The minimum distance required between two potential centroid points. This threshold acts as a second filter to ensure candidate centroids are adequately spaced apart.
- **Minimum weight score**: The minimum score a voxel must achieve to be considered as a potential centroid. This score is used towards the end of the clustering process to filter out less likely centroid candidates.

This explanation aims to provide a clear understanding of how the Fast Search algorithm identifies potential centroids through a series of hyperparameter-defined filters.

### Centers distribution comparison

| | Center 1 | Center 2 | Center 3 | Center 4 | Center 5 | Center 6 | Center 7 | Center 8 | Center 9 | Center 10 | Center 11 |
|---|---|---|---|---|---|---|---|---|---|---|---|
| **Center 1** | - | < * (46, 49) | < * (46, 45) | < * (46, 46) | < * (46, 9) | < * (46, 13) | < * (46, 71) | < * (46, 164) | < * (46, 20) | < * (46, 20) | < * (46, 22) |
| **Center 2** | < * (49, 46) | - | < * (49, 45) | < * (49, 46) | < * (49, 9) | < * (49, 13) | < * (49, 71) | < * (49, 164) | < * (49, 20) | < * (49, 20) | < * (49, 22) |
| **Center 3** | < * (45, 46) | < * (45, 49) | - | < * (45, 46) | < * (45, 9) | < * (45, 13) | < * (45, 71) | < * (45, 164) | < * (45, 20) | < * (45, 20) | < * (45, 22) |
| **Center 4** | < * (46, 46) | < * (46, 49) | < * (46, 45) | - | < * (46, 9) | < * (46, 13) | < * (46, 71) | < * (46, 164) | < * (46, 20) | < * (46, 20) | < * (46, 22) |
| **Center 5** | < * (9, 46) | < * (9, 49) | < * (9, 45) | < * (9, 46) | - | < * (9, 13) | < * (9, 71) | < * (9, 164) | < * (9, 20) | < * (9, 20) | < * (9, 22) |
| **Center 6** | < * (13, 46) | < * (13, 49) | < * (13, 45) | < * (13, 46) | < * (13, 9) | - | < * (9, 71) | < * (9, 164) | < * (9, 20) | < * (9, 20) | < * (9, 22) |
| **Center 7** | < * (71, 46) | < * (71, 49) | < * (71, 45) | < * (71, 46) | < * (71, 9) | < * (71, 13) | - | < * (71, 164) | < * (71, 20) | < * (71, 20) | < * (71, 22) |
| **Center 8** | < * (164, 46) | < * (164, 49) | < * (164, 45) | < * (164, 46) | < * (164, 9) | < * (164, 13) | < * (164, 71) | - | < * (164, 20) | < * (164, 20) | < * (164, 22) |
| **Center 9** | < * (20, 46) | < * (20, 49) | < * (20, 45) | < * (20, 46) | < * (20, 9) | < * (20, 13) | < * (20, 71) | < * (20, 164) | - | < * (20, 20) | < * (20, 22) |
| **Center 10** | < * (20, 46) | < * (20, 49) | < * (20, 45) | < * (20, 46) | < * (20, 9) | < * (20, 13) | < * (20, 71) | < * (20, 164) | < * (20, 20) | - | < * (20, 22) |
| **Center 11** | < * (22, 46) | < * (22, 49) | < * (22, 45) | < * (22, 46) | < * (22, 9) | < * (22, 13) | < * (22, 71) | < * (22, 164) | < * (22, 20) | < * (22, 20) | - |

**Table 2.** **Comparative Analysis of Center Distributions:** This table presents a comparative analysis of Adjusted P-values for volume Hounsfield Units distributions across different centers. Each cell details the Adjusted P-value for inter-center comparisons, with the respective sample sizes for the row and column centers presented underneath the P-value symbols. The notation '< *' signifies that both the P-value and Adjusted P-value (corrected using the Benjamini-Hochberg procedure for multiple comparisons) are below the threshold of 0.0001. These results indicate significant distributional disparities in volume Hounsfield Units across all examined centers.

### Ablation Study results on Internal Validation Set and Internal Test Set

| Dataset | $DSC_{seg}$ (%) | $IoU_{seg}$ (%) | $DSC_{bound}$ (%) | $HD95_{bound}$ (mm) | $Chamfer_{bound}$ (mm) |
|---|---|---|---|---|---|
| **Sinus** | | | | | |
| Internal Validation Set | 98.32 (97.94-98.71) | 96.79 (96.12-97.51) | 84.61 (81.70-87.71) | 0.68 (0.57-0.79) | 0.19 (0.17-0.22) |
| Internal Test Set | 97.17 (96.56-97.63) | 94.79 (93.91-95.60) | 78.02 (75.86-80.16) | 1.06 (0.81-1.37) | 0.29 (0.23-0.35) |
| **Inferior Alveolar Nerve** | | | | | |
| Internal Validation Set | 85.73 (83.66-87.77) | 78.48 (75.96-81.12) | 77.10 (74.46-80.03) | 5.34 (3.50-7.38) | 0.61 (0.46-0.78) |
| Internal Test Set | 85.50 (83.78-87.05) | 78.65 (76.78-80.49) | 78.22 (76.40-79.94) | 4.929 (3.890-5.976) | 0.63 (0.50-0.77) |

**Table 3. Ablation Study results on Internal Validation Set and Internal Test Set for the SISTR method:** this table presents the metrics of our SISTR method for Sinus and Inferior Alveolar Nerve segmentation on the Internal Validation Set and the Internal Test Set.

### Model architectures

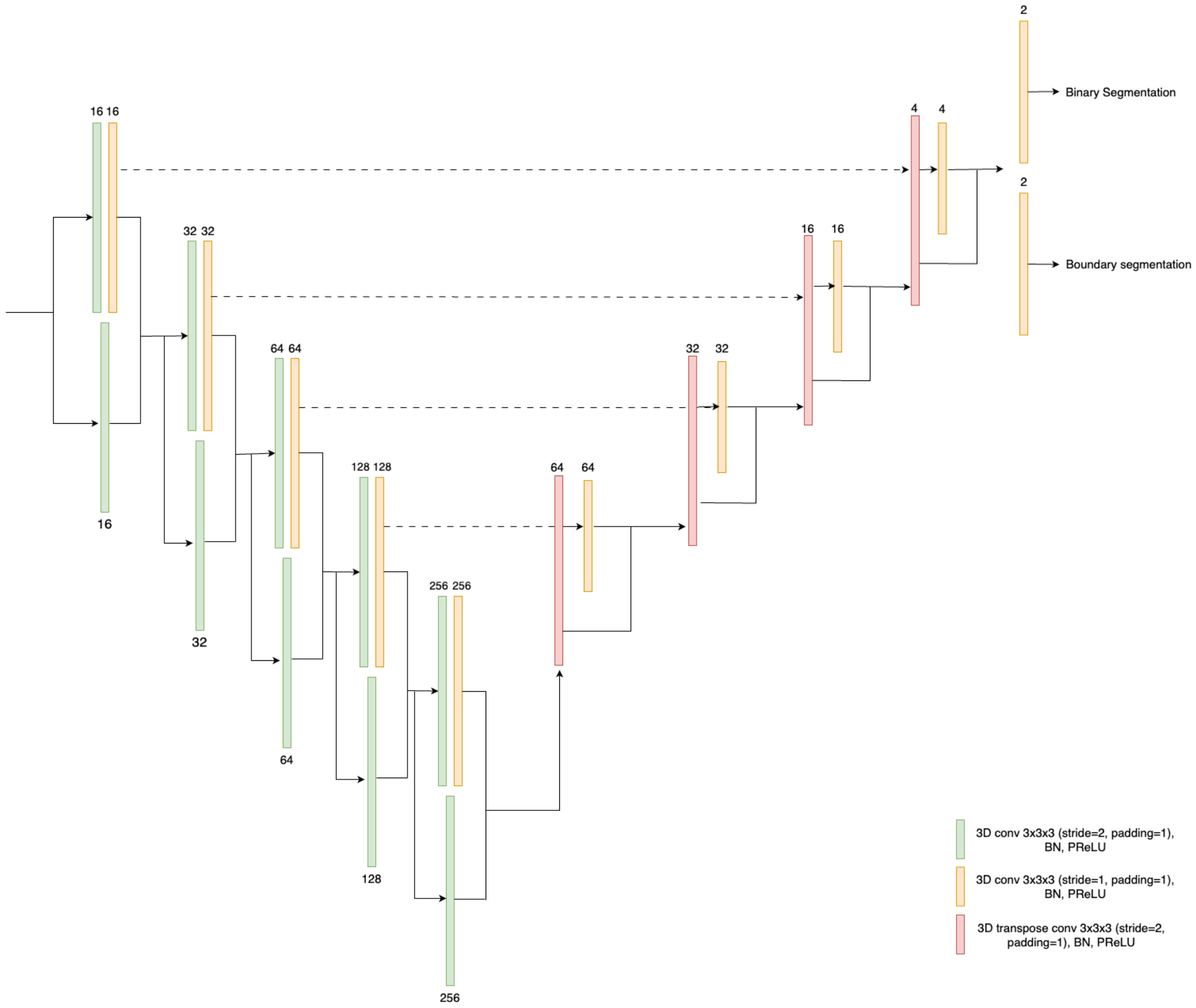


**Figure 1.** Network Architecture of the segmentation and boundary prediction model.

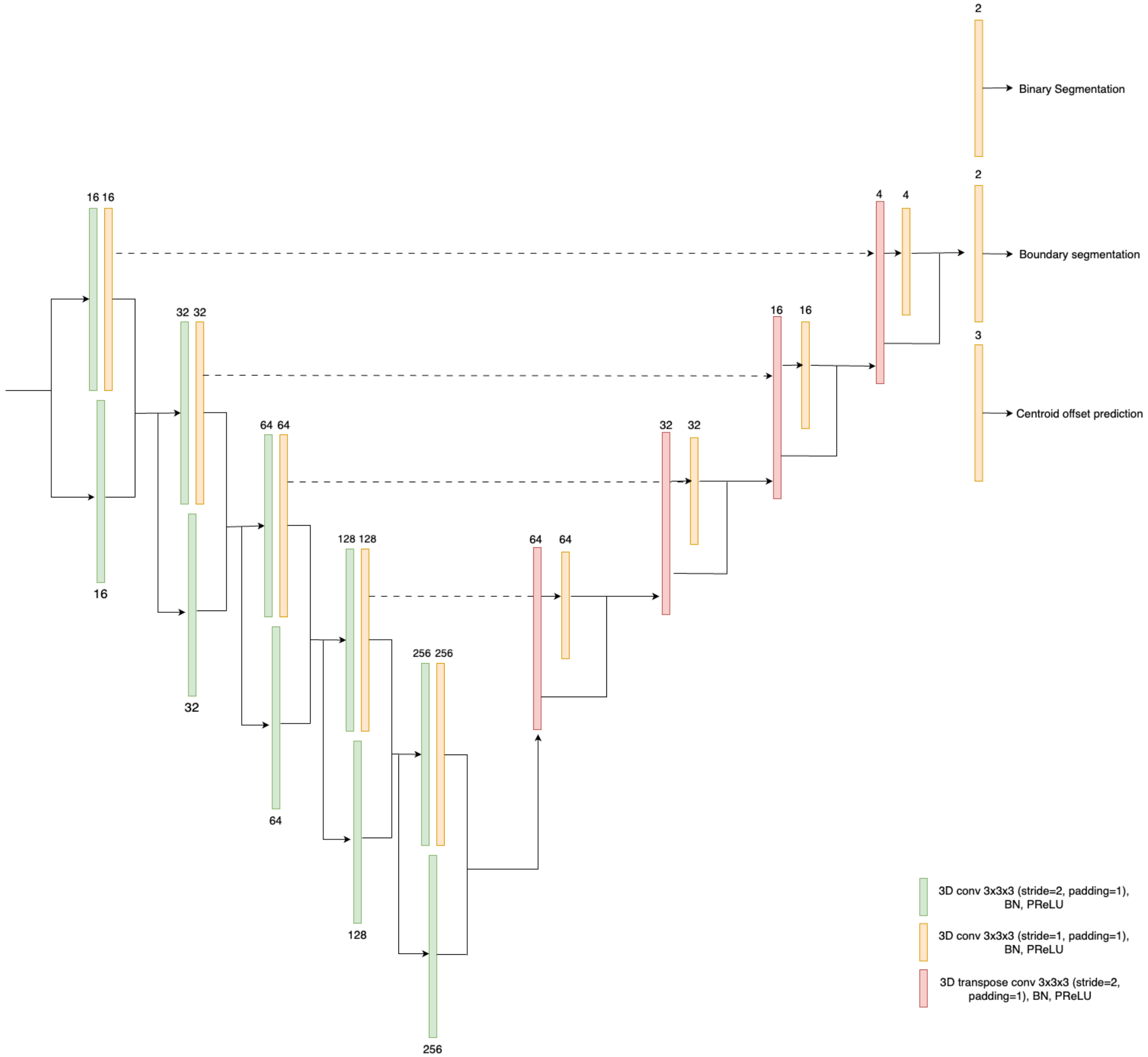


**Figure 2.** Network Architecture of the segmentation, boundary and offset model.

### Loss hyperparameters

Loss coefficients $\alpha_{bd}, \alpha_{seg}, \alpha_{off} \in \mathbb{R}^3$ in Equation (1) of the main manuscript were selected to balance the weights between losses, with the following coefficients:

- $\alpha_{bd} = 1$ for experiments with boundary prediction branch otherwise $\alpha_{bd} = 0$.
- $\alpha_{seg} = 1$ for all experiments.
- $\alpha_{off} = 0.33$ for all experiments.
